# Use of surgical antimicrobial prophylaxis and magnitude of surgical site infections among surgical patients at Hiwot Fana Comprehensive Specialized Hospital, Harar, Ethiopia

**DOI:** 10.1101/2025.11.06.25339731

**Authors:** Simon Tadesse, Badhaasaa Beyene, Elias Sertse

**Affiliations:** School of Medicine, College of Health and Medical Sciences, Haramaya University, Harar, Ethiopia

**Keywords:** Antibiotics, surgical care, comorbidity, wound classes

## Abstract

**Background:** Surgical site infections (SSIs) remain a common complication after surgery, particularly in low-resource settings. This study assessed the use of surgical antimicrobial prophylaxis (SAP) and the magnitude of SSI among Surgical patients at Hiwot Fana Comprehensive Specialised Hospital, Harar, Ethiopia.

**Methods:** A hospital-based retrospective chart review was conducted among 297 randomly selected patient records from November 2022 to October 2023. Data were analyzed using descriptive statistics and logistic regression to identify factors associated wth SSI.

**Results:** SAP was administered to 96 % of patients, predominantly ceftriaxone (72%). The overall SSI rate was 10.9 % (95% CI: 7.1-14.1). Factors significantly associated with SSI included comorbidity, previous surgery, ASA score ≥2, emergency procedures, contaminated/dirty wounds, and lack of prophylaxis.

**Conclusion:** Despite high SAP utilization, SSI prevalence remains moderate. Strengthening adherence to surgical antibiotic prophylaxis guidelines and optimizing timing and antibiotic choice are recommended to further reduce SSIs.

## Introduction

Surgical site infections are a common complication after surgery, posing a significant threat to patient health and increasing healthcare costs. Surgical antimicrobial prophylaxis involves administering antibiotics before, during, or after surgery to reduce the risk of infection at the surgical site.^1^ By targeting specific bacteria likely to contaminate the surgical area, SAP plays a crucial role in protecting patients.^2^ The choice of antibiotic in SAP depends on several factors, including: Type of surgery, Anatomical location, and Local antibiotic resistance patterns. Studies show that properly administered SAP can prevent 40-60% of SSIs.^3,4^ This translates to improved patient outcomes due to reduced risk of infections, shorter hospital stays, and faster recovery, and Lower healthcare costs.

Several factors increase a patient’s susceptibility to SSIs, including: general health, as in the case of underlying medical conditions like diabetes and obesity, the immune system can be weakened, surgery type: since High-contamination procedures pose a greater risk of infection, and Patient factors: Age, gender, and nutritional status can also influence infection risk.^5^

When used appropriately, SAP is a valuable tool for preventing SSIs, especially in high-risk patients. Effective SAP requires selecting the right antibiotic, administering in proper dosage and timing, and maintaining drug levels during long procedures.^6^ By promoting proper SAP use and awareness of risk factors, the burden of SSIs can be reduced, and patient outcomes after surgery can be improved. Despite national and WHO recommendations, inconsistent SAP practice persists across Ethiopian hospitals, contributing to variable SSI rates. This study provides local data to inform evidence-based surgical infection prevention strategies.^7^

## Material and methods

### Study period and setting

The study, i.e, chart review, was conducted from November 25, 2022, to November 30, 2022, at Hiwot Fana Comprehensive Specialized Hospital (HFCSH) located in Harari region, which is 526km from the capital city, Addis Ababa to the Eastern part of Ethiopia. Hiwot Fana Comprehensive Specialized Hospital serves as a teaching university hospital and referral center for eastern Hararge, part of western Hararge, and the nearby Somali region. HFCSH offers multispecialty services that encompass various surgical specialities ^8^

Study design: An institution-based cross-sectional study design was employed.

Source populations: Patients who are admitted to the surgical unit and undergo surgery for elective or emergency surgery in Hiwot Fana Specialized University Hospital

Study populations: Patients who are admitted to the surgical unit and undergo surgery for elective or emergency surgery in Hiwot Fana Comprehensive Specialized Hospital

### Eligibility Criteria

#### Inclusion Criteria

Medical charts of patients who were admitted and operated at Hiwot Fana Comprehensive Specialized Hospital during November 1, 2022 – October 31, 2023 were included.

#### Exclusion Criteria

Patients with incomplete medical charts, those self-discharged patients, and patients transferred to other facilities were excluded.

#### Sample Size Determination

For the prevalence of SSI, the single population proportion formula was used and computed by considering the prevalence of 19.6%.^9^, 5% precision (d=0.05), 95% level of confidence (z=1.96), and 10% non-response rate.

The sample size was calculated as follows:

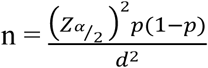

#### Where: n=sample size required

Zα/2 = the standard normal deviation corresponding to 95% confidence level = 1.96

#### P: prevalence of SSI

#### d: margin of error (half of the degrees of freedom)

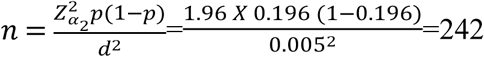

Then, a 10% non-response rate was added, and the final sample size became 266.

Similarly, the single population proportion formula is used for calculating the sample size for the second objective. According to Argaw et al, the proportion of antibiotic utilization among surgical patients was 41%.^10^

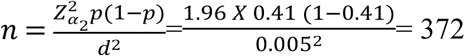

As the source population is less than 10000, we used the finite population correction formula.

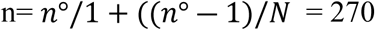

Where: n=corrected Sample size,

#### n°= sample size calculated early (372) and

N= total number of the population in the study area (1000).

Finally, after adding a 10% non-response proportion, a total of 297 study participants were involved in the study.

A simple random sampling technique is used to select charts of surgical patients from the OR logbook. The source population is estimated from the twelve-month (November 1, 2022– October 31, 2023) report of the hospital. From the study population of 1000 patients, who underwent surgery during this period, 297 patients were selected using a lottery-type random sampling technique using a random number generator. If the selected patient had incomplete data or self-discharged, the chart of the patient immediately next to him/her was replaced.

### Data collection tool and procedure

The structured Data extraction checklist adopted from a review of different literature was used to collect data from patients’ charts about sociodemographic characteristics, clinically related information, and prophylactic antibiotic regimens. Two trained and experienced nurses were used for the data collection.

### Data quality control

The data collection tool was pretested on 5% of the sample size in Jugal General Hospital for its consistency, completeness, and ease for understanding. The data collectors were trained for one day before the data collection period. The data collection process was closely supervised by the principal investigator to ensure the completeness, accuracy, and consistency of data collection. The collected data was thoroughly checked for completeness daily by the principal investigator. When any gap is identified, it is communicated to data collectors daily.

### Variables

#### Dependent variable

Surgical site infection

#### Independent variables

Sociodemographic characteristics: Age and gender, Residence of patients

Clinical related information: type of surgery, comorbidity, preoperative ASA score, wound classification, duration of preoperative hospital stays, Length of Surgery.

Antimicrobial prophylaxis use.

#### Operational definition

**Surgical site infection**: Surgical site infections are infections that develop within 30 days after an operation or surveillance of surgical wound infection implementation within 90 days after surgery when an implant is placed ^11^

**Appropriate antimicrobial prophylaxis indication**: indication of a prophylactic antimicrobial agent that is consistent with the WHO SSI prevention guideline recommendation.^12^

**Clean wound**: elective, not emergency, nontraumatic, and primarily closed; no acute inflammation; no break in technique; respiratory, gastrointestinal, biliary, and genitourinary tracts not entered.^6^

**Clean contaminated wound**: urgent or emergency case that is otherwise clean; elective opening of respiratory, gastrointestinal, biliary, or genitourinary tract with minimal spillage (e.g., appendectomy) not encountering infected urine or bile; minor technique break ^6^

**Contaminated wound**: non-purulent inflammation; gross spillage from gastrointestinal tract; entry into biliary or genitourinary tract in the presence of infected bile or urine; major break in technique; penetrating trauma < 4 hours old; chronic open wounds to be grafted or covered. ^6^

### Data analysis methods

Data was checked, cleaned for any deficit before, and entered into Epi-Info 7 and then exported to SPSS version 26.0 for analysis. Data was analyzed with both descriptive analyses, such as frequencies and percentages, and inferential analysis was also conducted with chi chi-square test and Logistic regression analysis. The assumption for binary logistic regression includes: the goodness of fit was checked by the Hosmer-Lemeshow statistic and omnibus tests. Multi-collinearity test was carried out to see the correlation between independent variables by using the standard error and collinearity statistics (Variance inflation factors > 10 and standard error > 2 considered as suggestive of the existence of multi-collinearity). For measuring the strength of the association between the outcome and independent variables, Crude Odds Ratio (COR) and Adjusted Odds Ratio (AOR) along with 95% Confidence interval (CI) were calculated. All variables with a p-value of < 0.25 were further analyzed with multivariate logistic regression analysis to control for confounders. The Association was considered Significant at a 95% confidence interval and p-value set at less than 5%, using SPSS, version 26.0.

### Ethical considerations

Ethical clearance was obtained from Haramaya University Institutional Health Research Ethics Review Committee (Ref. No. IHRERC/224/2023). Then, an official letter for cooperation and permission was delivered to Hiwot Fana Comprehensive Specialized Hospital administration and all responsible bodies to get permission for the study. Moreover, a brief explanation was given to the hospital manager concerning the purpose of the study and the procedures used to collect the data. Then, an information sheet and an informed, voluntary, written, and signed consent form for the Head of the Hospital were given. After getting permission, patients’ charts that fulfilled the criteria were selected. The confidentiality of the data collected is maintained; patient names were not included during data collection, and authors had no access to information that could identify individual participants during or after data collection

## Results

### Socio-demographic and clinical characteristics

Among 297 surgical patients, 69.7% patients were males, and 75.4% resided in rural areas. Most surgeries (53.2%) were emergency procedures, and 70% were clean-contaminated wounds. Refer Table 1 for details.

**Table 1.** Socio-Demographic and Clinical Characteristics of the Study Participants (N = 297)

| Variables | Categories | N (%) |
| --- | --- | --- |
| Age | <20 | 66 (22.2%) |
|  | 21-40 | 130 (43.8) |
|  | 41-60 | 66(22.2%) |
|  | >60 | 35(11.8%) |
| Sex | Male | 207(69.7 %) |
|  | Female | 90 (30.3%) |
| Residence | Rural | 224(75.4%) |
|  | Urban | 73(24.6%) |
| Comorbidity | Yes | 28 (9.4%) |
|  | No | 269(90.6%) |
| Pre operative ASA score | I | 200(67.3 %) |
|  | II | 66 (22.2 %) |
|  | III | 25 (8.4 %) |
|  | IV | 6 (2.0 %) |
| Previous surgery history | Yes | 22 (7.4 %) |
|  | No | 275 (92.6 %) |
| Types of surgery | Emergency | 158 (53.2%) |
|  | Elective | 139 (46.8 %) |
| Preoperative hospital stay | <72 hours | 215 (72.4 %) |
|  | 72hrs-1 week | 64 (21.5 %) |
|  | >1 week | 18 (6.1 %) |
| Duration of surgery (in hours) | <1 hour | 109 (36.7 %) |
|  | 1-2 hours | 144 (48.5 %) |
|  | >2 hours | 44 (14.8 %) |
| Wound class | Clean | 58 (19.5 %) |
|  | Clean- contaminated | 208 (70 %) |
|  | Contaminated & Dirty | 31 (10.5%) |

On the practice of surgical antimicrobial prophylaxis: Two hundred eighty-five (96%) of patients had received surgical antimicrobial prophylaxis, with ceftriaxone being the most frequently used (72%) antimicrobial. The intravenous route was used in 95.3 % of patients. For 222 (74.7%) patients, and antimicrobials were administered within 60 minutes before the skin incision. See Table 2.

**Table 2.**
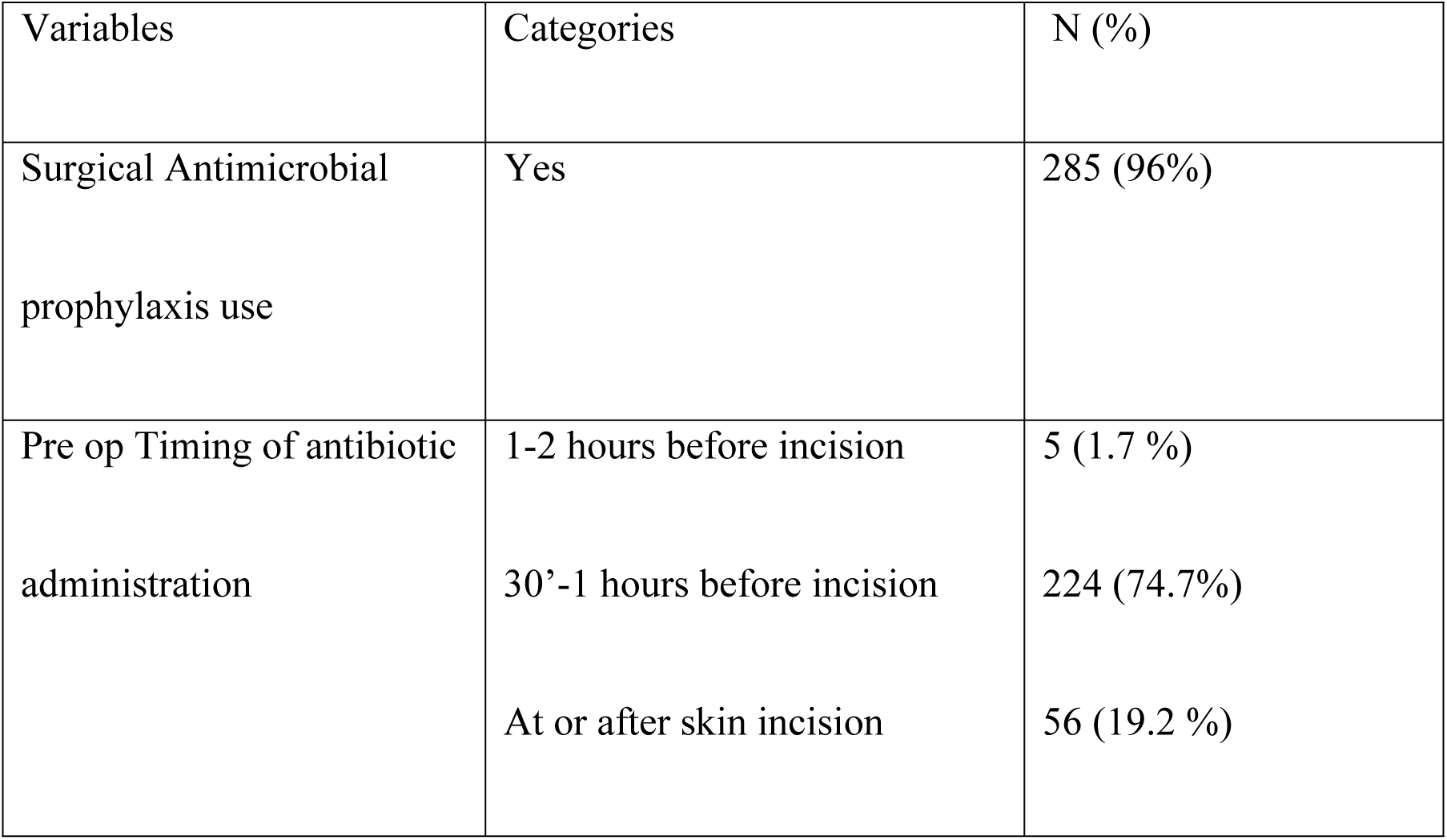
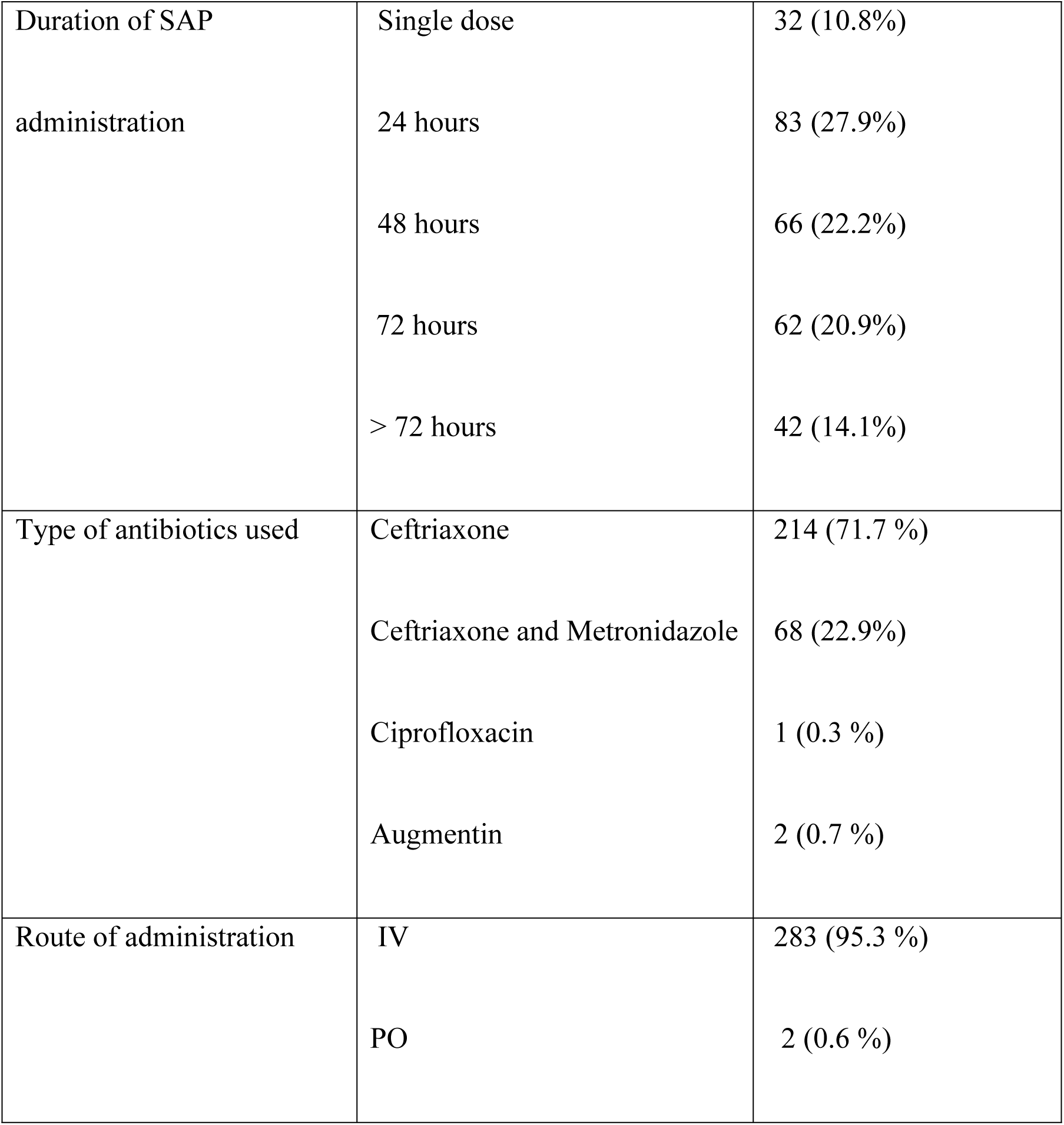
Practice of Surgical antimicrobial prophylaxis.

### Magnitude of surgical site infections and associated factors

Finally, from this study, it was retrieved that the diagnosis of surgical site infection was made in 32 patients (10.9 %; 95% CI: 7.1-14.1), with 23 of them (71.8 %) having superficial SSI.

According to this study, bivariable logistic regression analysis of patients who developed SSI had significant association (with p value of < 0.25) with patients having comorbidity, age range greater than twenty, Rural residence, having preoperative ASA score two and three, previous history of Surgery, Emergency procedure, procedures lasting one to two hours, contaminated and Dirty wound classes and devoid of prophylaxis were taken as candidate for Multivariable analysis.

Upon multivariable logistic regression analysis, SSI was 3.6 times more likely to happen among patients who had a history of previous surgery. (AOR 3.36; 95%CI: 1.08-10.4), 4.8 more likely to happen among patients with a Preoperative ASA score of two (AOR 4.89; 95%CI:1.72-13.7) and 7.7 more likely to happen with an ASA score of three. (AOR 7.70; 95%CI: 1.80-32.8) respectively, And Patients undergoing emergency procedures were three times likely to have SSI than patients operated on an Elective basis. (AOR 3.14; 95%CI: 1.13-8.60). In addition, the odds of developing SSI were 2.85 times higher among patients with comorbidity than among patients without comorbidity. (AOR 2.85; 95%CI: 1.07 – 7.56) On the other hand, contaminated and dirty wound classes were fifteen times likely to develop SSI than clean wound types. (AOR 15.3; 95%CI: 2.66-88.9). Patients who did not get prophylactic antibiotics were six (5.8) times more likely to develop SSI than patients who had prophylactic antibiotics. (AOR 5.88; 95%CI: 1.88-18.1), whereas Age, rural residence, and duration of procedure were dropped. Table 3

**Table 3.**
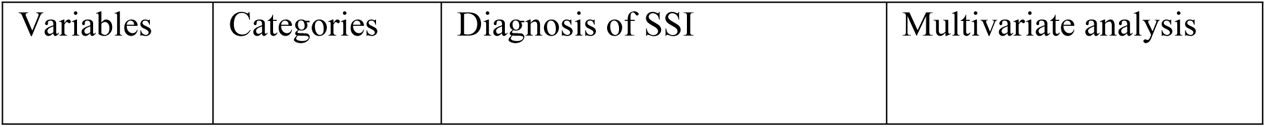
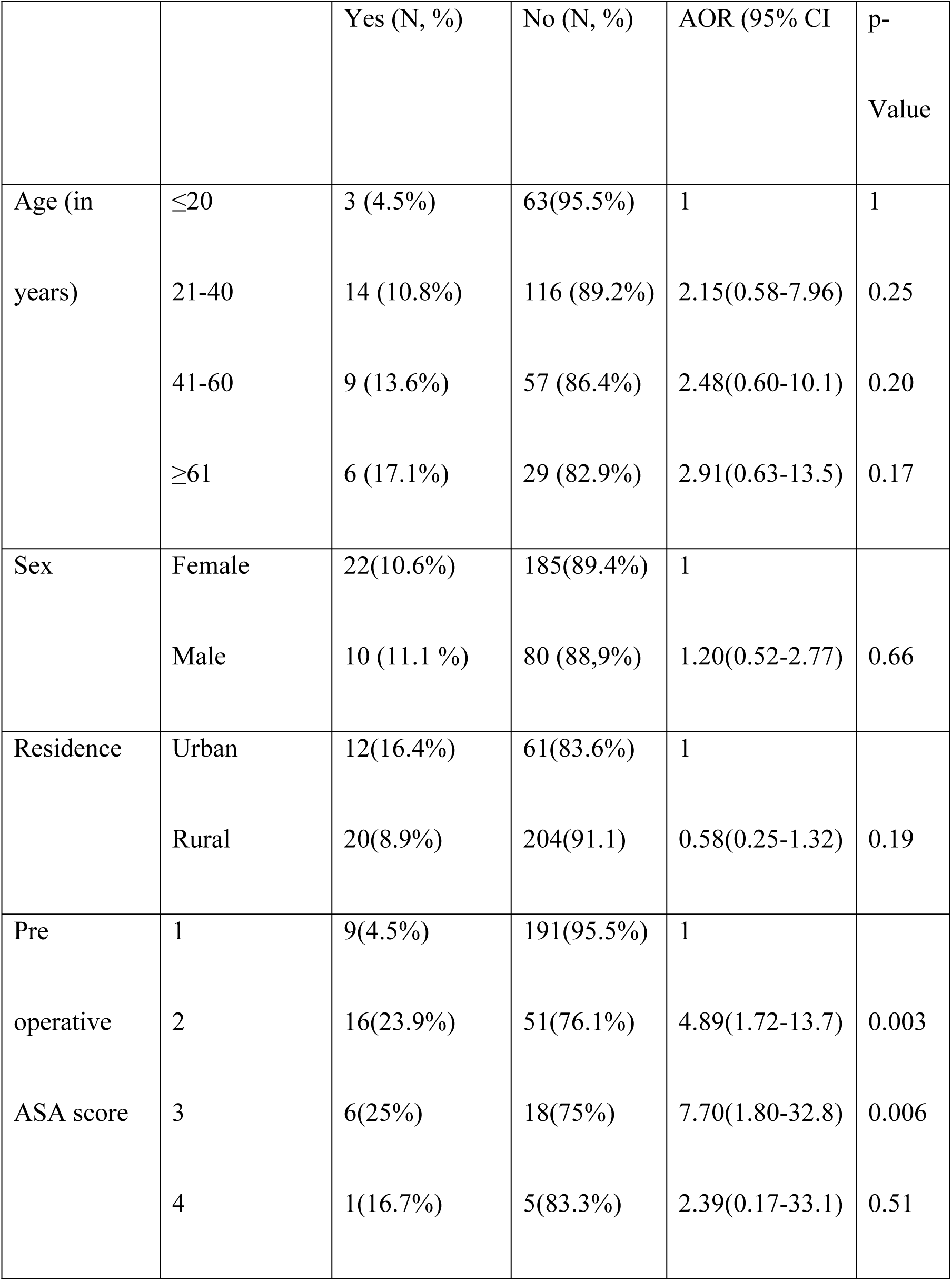
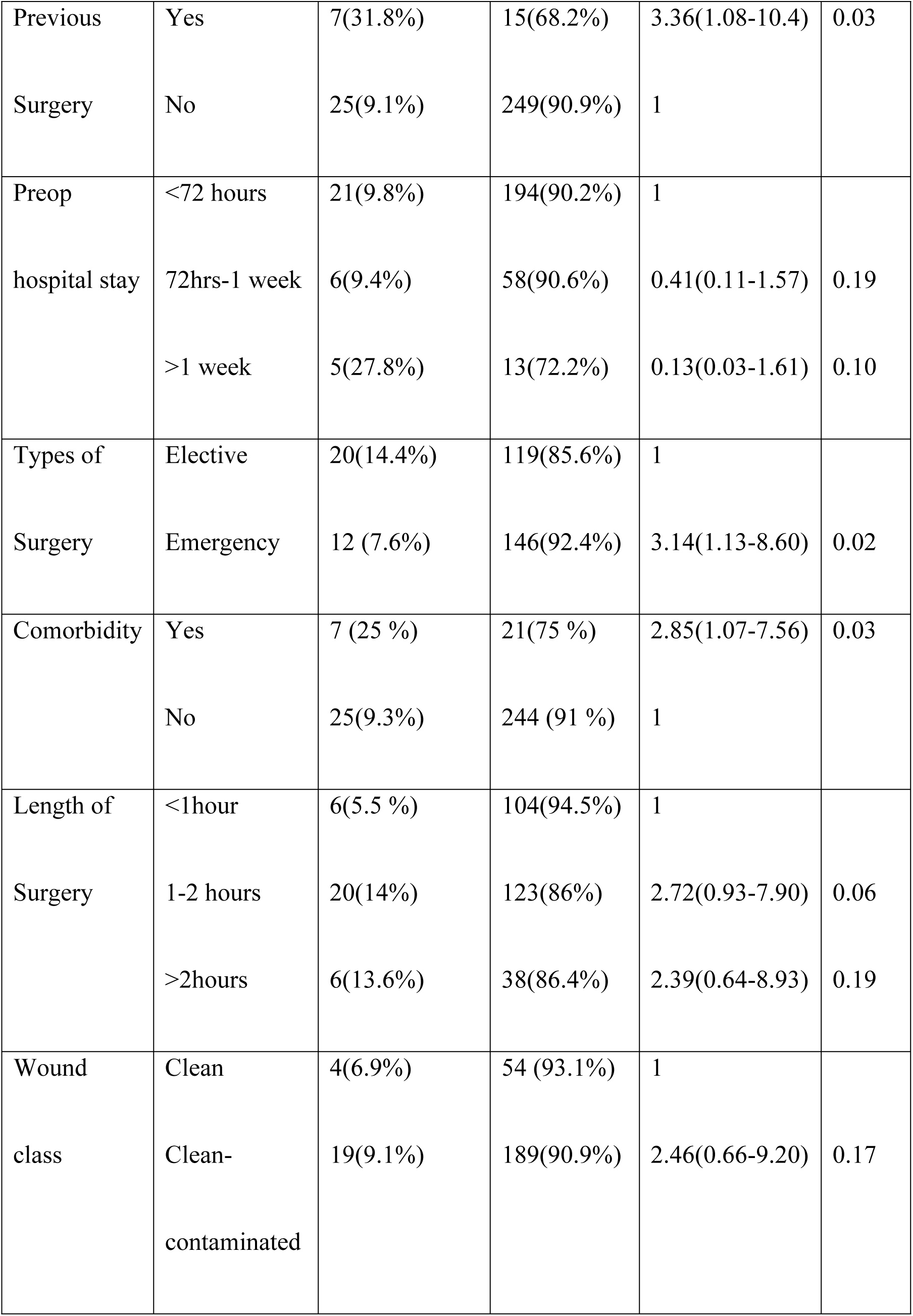
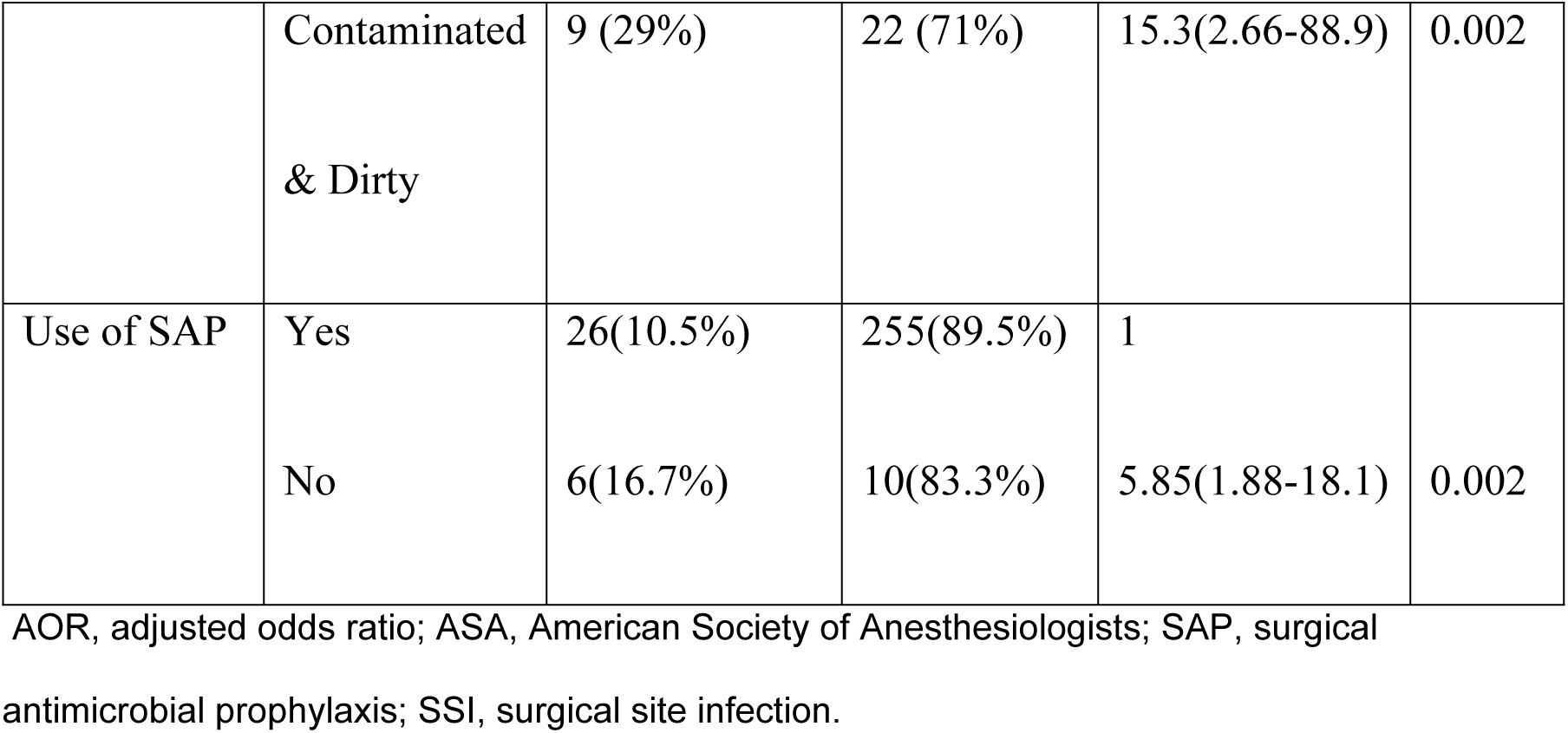
Multivariable analysis of Factors associated with Surgical site infections among Surgical patients.

## Discussion

In this study, the use of antibiotics for surgical prophylaxis purposes and the magnitude of SSI were assessed in Hiwot Fana Comprehensive Specialized Hospital, Harar, Ethiopia. Sociodemographic and clinical characteristics, Surgical antimicrobial prophylaxis utilization practice in surgery, magnitude of SSIs were studied.

Surgical site infections were observed in 10.9% patients. The prevalence of SSI is comparable to studies done in SPHMMC (11.1 %)^10^, Fisha et al (9.9%).^13^ However, the prevalence was lower than studies conducted by Misganaw et al(23.4%)^14^ and Mezemir et al (24.6%).^15^ In addition, it was lower than the recent systematic review performed in Ethiopian literature (25.22%).^16^ This may be attributed to better practice of prophylactic surgical antibiotic use (96 %) in our circumstance.

More over 71.8% of SSIs were superficial followed by organ space (15.6%), and Deep (12.5%) infections which is different from Mezemir et al,^15^ study that reported 40.7% and 37.4% were deep and organ space infections, respectively: the finding is also different to a lesser extent from a study by Kefale et al,^9^ which states 14.7 % Superficial SSI followed by 3 % deep SSI.

In the present study, more than half (53.2%) of surgeries were emergency which is higher than the study from kefala et al (46.6%).^9^ And, most of types of surgical wound in our study was clean and clean contaminated with combined percentage of the remaining contaminated and dirty wound being 10.5 % which is significantly lower than 36% from the study by kefale et al. ^9^ regarding duration of surgery also majority of them (85.2%), last less than 2 hours which is higher than 71.1% from kefala et al. ^9^

As to prophylactic antibiotic use 96 % of surgical patients got Surgical antibiotic prophylaxis. which is greater than, Alamrew et al (72.1%),^11^ Kefale et al (88.6%)^9^ and Lijaemiro et al (92%)^17^ and lower than Ayele and Taye study which is 100 % utilization rate. ^18^ The difference may be based on the variation of cases and adherence of professionals involved to preoperative guidelines. International guidelines promote the use of less costly and narrow spectrum antibiotics for prophylaxis; Cefazolin is widely indicated as first choice for most of surgical procedure. In our study however, 71.7% of subjects were managed by ceftriaxone. Although this study is in line with study done in Addis Ababa where 70% of procedures were managed by Ceftriaxone;^10^ it is in contrast to Botswana ^19^ and Sudanese^20^ studies in which they reported Cefotaxime and Cefazolin as the most commonly used prophylactic agent respectively. According to American Family Physicians recommendations, prophylactic antibiotics should be initiated within 1 hour before surgical incision.^21^ This supports our study finding as most of our study participants (222, 74.7%) received SAP within 1 hour before surgical incision.

In this study few Risk factors were identified which are related with SSI. Patients undergoing emergency procedures were three times likely to have SSI than patients operated on Elective basis. This is justifiable in that emergency procedures will be done without adequate preoperative preparation and measures to reduce the microbial load in the operative field therefore contamination of the surgical site can be highly likely.

In addition, the odds of developing SSI were 2.85 times higher among patients with comorbidity than patients without comorbidity. This is in line with Misganaw et al^14^ study. The presence of comorbidity may expose patients to infection due to lowered immunity.

Additionally, history of previous surgery, poses three times more risk of SSI for those who had it. This is in line with Tamrat et al study.^22^ Having previous surgery and scar can lead to difficult surgery and also increased duration of procedures which can lead to increased contamination of operative field; additionally stress of prolonged anesthesia, possibility of increased blood loss can also decrease the tissue level of the surgical antimicrobial prophylaxis which, with its final effect increases the risk of SSIs.

On the other hand, Preoperative ASA score two and three were four and seven times likely to develop SSI than ASA score 1 patients, this is in line with Shiferaw et al study;^23^ global guidelines for the prevention of surgical site infection also supported that an ASA score of at least 3 recognized as factor associated with an increased risk of SSI,^24^ as impaired physical activity will affect level of Immunity.

Contaminated and dirty wound classes together were fifteen times likely to develop SSI than clean wound types. This finding is in line with Weldu et al,^25^ and Misganaw et al^14^ studies. These types of wound classes are suitable for the colonization and multiplication of different pathogens and surgeries with an increased microbial load in the operative field are associated with higher risk of SSI. In addition, evidence support that, treating infections on the operating site, if possible, or postponing the surgery until the infection has cleared will decrease the risk of SSI.

Finally, Patients who did not get prophylactic antibiotics were six (5.8) times more likely to develop SSI than patients who had prophylactic antibiotics. which is in line with Misganaw et al^14^ and Alamrew et al^11^ studies. Administering prophylactic antibiotic prevent colonization and spread of microbes which will decrease the risk of Surgical site infection.

### Limitation of the study

The retrospective study design, in ability to confirm microbiological data, single institution scope of the study and possible documentation bias are the limitations of the study.

## Conclusion

This study showed that the prevalence of Surgical site infection was moderate as compared to other Ethiopian studies and there is high rate of utilization of Surgical antimicrobial prophylaxis. Emergency Surgical procedures, presence of comorbidity, contaminated and Dirty wound classes and devoid of prophylactic antibiotics were significantly associated with Surgical site infection.

Despite high SAP use, inappropriate antibiotic selection and timing may contribute to the residual infections. Focused interventions and periodic audits are recommended.

## Data Availability

All relevant data are within the manuscript and its Supporting Information files.

## Acknowledgments

The authors thank Haramaya University for ethical clearance and Hiwot Fana Comprehensive Specialised Hospital for facilitating data collection.

## Data Availability

All relevant data are within the manuscript and its Supporting Information files.

